# Adverse clinical outcomes in people at clinical high-risk for psychosis related to activity and glutamate function in the hippocampus

**DOI:** 10.1101/2021.03.19.21253942

**Authors:** Paul Allen, Emily J. Hird, Natasza Orlov, Gemma Modinos, Matthijs Bossong, Mathilde Antoniades, Carly Sampson, Matilda Azis, Oliver Howes, James Stone, Jesus Perez, Matthew Broome, Antony A. Grace, Philip McGuire

## Abstract

Preclinical models suggest that psychosis involves alterations in activity and glutamate function in the hippocampus, driving dopamine activity through projections to the striatum. The extent to which this model applies to the onset of psychosis in clinical subjects is unclear. We assessed whether interactions between hippocampal glutamatergic function and activity/striatal-connectivity are associated with adverse clinical outcomes in people at clinical high-risk (CHR) for psychosis. We measured functional Magnetic Resonance Imaging of hippocampal activation/connectivity, and ^1^H-Magnetic Resonance Spectroscopy of hippocampal glutamatergic metabolites in 75 CHR participants and 31 healthy volunteers. At follow-up, 12 CHR participants had transitioned to psychosis and 63 had not. Within the clinical high-risk cohort, at follow-up, 35 and 17 participants had a poor or a good functional outcome, respectively. The onset of psychosis (*p*_*peak*FWE_ =.003, t=4.4, z=4.19) and a poor functional outcome (*p*_*peak*FWE_ <.001, t=5.52, z=4.81 and *p*_*peak*FWE_ <.001, t=5.25, z=4.62) were associated with a negative correlation between hippocampal activation and hippocampal Glx concentration at baseline. In addition, there was a negative association between hippocampal Glx concentration and hippocampo-striatal connectivity (*p*_*peak*FWE_ *=*.016, t=3.73, z=3.39, *p*_*peak*FWE_ *=*.014, t=3.78, z=3.42, *p*_*peak*FWE_ *=*.011, t=4.45, z=3.91, *p*_*peak*FWE_ *=*.003, t=4.92, z=4.23) in the total CHR sample, not seen in healthy volunteers. As predicted by preclinical models, adverse clinical outcomes in people at risk for psychosis are associated with altered interactions between hippocampal activity and glutamatergic function.

## Introduction

The onset of psychosis is commonly preceded by a clinical high-risk (CHR) phase, characterised by ‘attenuated’ psychotic symptoms and a marked decline in social and occupational functioning ^1^. This syndrome is associated with a 20-30% risk of developing psychosis in the following 2-3 years ^1–3^. Data from preclinical studies in rats, and in schizophrenia patients, suggest that the onset of psychosis involves an increase in resting hippocampal activity ^4–6^, which may be secondary to a dysregulation of hippocampal glutamatergic neurotransmission ^5,7^. This primary hippocampal dysfunction is then thought to drive an increase in subcortical dopamine activity, through modulatory glutamatergic projections from hippocampus to striatum ^6^ (figure 1).

**Figure 1:**
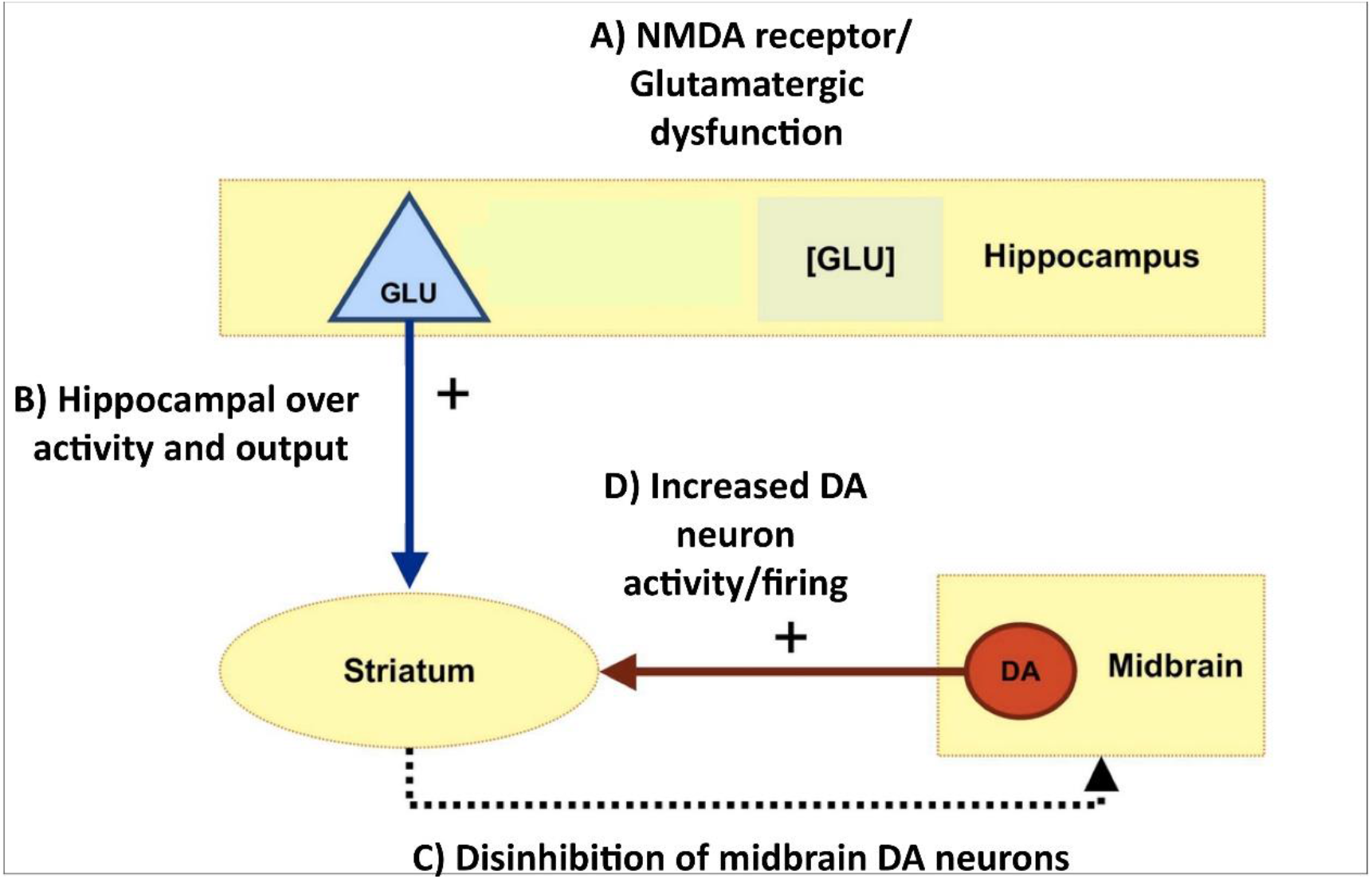
Preclinical model hippocampal – striatal-midbrain circuit A) Hippocampal glutamatergic function B) drives hippocampal activity and output to the striatum that C & D) deregulates striatal dopamine function in psychosis.

Neuroimaging studies in CHR populations indicate that the CHR state is associated with increased resting activity and perfusion in the hippocampus^8–10^, and with altered hippocampal activation in response to tasks that involve salience processing ^11–13^ or verbal memory ^14^. Parallel work suggests that the concentration of glutamatergic metabolites in the hippocampus is also altered in CHR subjects ^15^ and is related to hippocampal activation during verbal memory tasks in this group ^16^. Finally, studies of functional connectivity between the hippocampus and the striatum indicate that this is altered in CHR individuals ^12,13^. Some of these neuroimaging findings in CHR samples have been linked to adverse clinical outcomes subsequent to scanning. For example, alterations in hippocampal activation ^17^ and glutamate levels ^15^ have each been independently linked to the later onset of psychosis.

To date, associations between neuroimaging measures and clinical outcomes in CHR samples have largely been identified in studies of a single neuroimaging metric. However, contemporary models emphasise the interrelated nature of physiological and neurochemical dysfunction in the hippocampus, and its interaction with the striatum through descending glutamatergic connections. Given this aetiological complexity, assessing *interactions* between neuroimaging measures of different abnormalities might provide a useful test of preclinical predictions in humans, and better predict clinical outcomes in CHR subjects than a single measure.

The aim of the present case-control study was to use multi-modal neuroimaging data to examine whether clinical outcomes in CHR participants are influenced by interactions between hippocampal activity, function, and hippocampo-striatal connectivity. We tested the hypothesis that alterations in the relationship between these measures would be associated with adverse subsequent clinical outcomes.

## Methods

### Participants

One hundred and sixteen individuals were recruited to the study. 85 participants were at clinical high risk of psychosis (CHR), and 31 were healthy controls (HC). The study was approved by the National Research Ethics Service Committee of London-Camberwell St Giles, United Kingdom. All participants gave written informed consent. Data collection took place between November 1, 2011 and November 1, 2017. Clinical high-risk (CHR) participants were recruited through four early detection services for people at clinical high risk for psychosis: Outreach and Support in South London (OASIS), the West London Early Intervention service, the Cambridge Early Onset service (CAMEO), and the Coventry and Warwickshire Partnership NHS trust. CHR participants were assessed using the Comprehensive Assessment of At Risk Mental States (CAARMS) ^1,2^. Individuals were excluded from the CHR group based on the following criteria: past/present diagnosis of psychotic disorders, past/present familiar history of neurological illness, substance abuse/dependence as defined using DSM-5 criteria ^3^, or contraindication to MRI scanning.

Healthy control (HC) participants were recruited from the same geographical locations as CHR participants. HC participants were native English speakers, did not have a personal or familial history of psychiatric or neurological disorder and were not using prescription medication as assessed via self-report. Further exclusion criteria were self-reported illicit substance use in the week before MRI scanning or alcohol use in the 24 hours before MRI scanning.

Premorbid IQ was assessed using the National Adult Reading Test (NART) ^6^, and handedness using the Annett Handedness Scale ^7^. Participants self-reported information on gender, tobacco use (number of cigarettes smoked per day) and cannabis use (0 indicated no use; 1 indicated experimental use; 2 indicated occasional use; 3 indicated moderate use; 4 indicated severe use).

The main outcome measures of the study were hippocampal ^1^H-MRS glutamatergic and fMRI data acquired from the same participants during the same MRI scanning sessions. These two datasets have previously been analysed and reported separately ^13,15^. The current study included all participants where both ^1^H-MRS and fMRI data were available. As such, the final sample comprised 75 CHR participants and 31 HCs. See Table 1 for reported demographic and clinical outcome characteristics.

**Table 1:** Demographic, clinical and medication data at presentation, divided by group and outcome.

| Measure at presentation | HCs vs. CHR |  |  | CHR-T vs. CHR-NT |  |  | CHR-good vs. CHR-poor |  |  |
| --- | --- | --- | --- | --- | --- | --- | --- | --- | --- |
|  | HC (N=31) | CHR (N=75) | P | CHR-NTR (N=63) | CHR-TR (N=12) | P | CHR-good (N=17) | CHR-poor (N=35) | P |
| Age in years, mean (SD) | 24.96<br>(4.06) | 22.62<br>(3.63) | .01 | 22.78<br>(3.78) | 21.83<br>(2.69) | .31 | 22.24<br>(3.38) | 22.89<br>(3.61) | .53 |
| NART IQ, mean (SD) | 104.86<br>(13.66) | 103.43<br>(14.67) | .65 | 103.55<br>(15.61) | 102.82<br>(8.58) | .83 | 107.38<br>(9.22) | 104.42<br>(15.43) | .41 |
| Years in education, mean (SD) | 15.8<br>(3.35) | 14.58<br>(2.22) | .07 | 14.65<br>(2.2) | 14.18<br>(2.36) | .55 | 15<br>(2.39) | 14.26<br>(2.14) | .29 |
| Positive CAARMS, mean (SD) | N/A | 10.13<br>(4.08) | N/A | 9.95<br>(4.14) | 11.08<br>(3.75) | .36 | 9.88<br>(4.03) | 10.54<br>(4.38) | .59 |
| Negative CAARMS, mean (SD) | N/A | 5.47<br>(4.17) | N/A | 5.35<br>(4.09) | 6.09<br>(4.74) | .64 | 6.76<br>(3.95) | 5.26<br>(4.29) | .22 |
| Total CAARMS, mean (SD) | N/A | 43.57<br>(21.74) | N/A | 42.65<br>(22.24) | 49.3<br>(18.27) | .32 | 44.44<br>(19.66) | 43.35<br>(20.61) | .86 |
| GAF, mean (SD) | 92.85<br>(4.98) | 57.81<br>(9.49) | <.001 | 58.56<br>(9.71) | 54.08<br>(7.56) | .09 | 57.03<br>(9.42) | 55.27<br>(9.67) | .55 |
| HAM-A, mean (SD) | 3.42<br>(4.18) | 18.22<br>(11.26) | <.001 | 16.95<br>(10.54) | 25<br>(13.16) | .11 | 17.61<br>(13.49) | 20.27<br>(11.9) | .55 |
| HAM-D, mean (SD) | 1.66<br>(3.52) | 17.67<br>(11.15) | <.001 | 16.79<br>(11.26) | 22.33<br>(9.78) | .15 | 15.77<br>(10.79) | 19.15<br>(11.91) | .38 |
| Cigarettes per day, mean (SD) | 1.93<br>(3.36) | 6.32<br>(9.09) | .001 | 7.12<br>(9.56) | 2<br>(3.76) | .004 | 6.82<br>(10.63) | 6.36<br>(8.91) | .88 |
| Daily units of alcohol, mean (SD) | 1.66<br>(2.22) | 1.59<br>(3.36) | .91 | 1.72<br>(3.63) | 0.91<br>(.7) | .12 | 1.41<br>(1) | 1.61<br>(4.13) | .79 |
| Current Cannabis use, median (range) | 0<br>(0-3) | 0<br>(0-4) | .74 | 0<br>(4) | 0<br>(4) | .52 | 1.2<br>(1.6) | 0.9<br>(1.2) | .17 |
| Currently taking antipsychotic medication, no (%) | 0<br>(0) | 10<br>(13) | .03 | 9<br>(14.3) | 1<br>(8.3) | .58 | 3<br>(17.6) | 3<br>(8.6) | .07 |
| Female sex, no (%) | 16<br>(52) | 33<br>(44) | .42 | 27<br>(43) | 6<br>(50) | .65 | 8<br>(47.1) | 13<br>(37.1) | .49 |
| Right-handed, no (%) | 28<br>(90) | 63<br>(84) | .52 | 52<br>(83) | 11<br>(92) | .71 | 16<br>(94.1) | 27<br>(77.1) | .38 |

### Clinical measures

After recruitment, the following clinical measures were collected at King’s College London on the day of MRI scanning by trained assessors: psychopathology using the CAARMS ^18^; overall functioning using the Global Assessment of Function (GAF) ^19^, and anxiety and depression symptoms using the Hamilton Anxiety and Depression Scale (HAM-A/HAM-D)^20^.

### Clinical follow-up

CHR participants were followed-up at a mean of 18.4 months (SD=12.8 months) after MRI scanning to determine clinical and functional outcomes. Transition to psychosis was assessed using the CAARMS Psychosis Threshold criteria ^18^ and confirmed with the Structured Clinical Interview for Diagnosis ^21^, administered by a psychiatrist trained in its use. Of the 75 CHR individuals included in the current analysis, 23 were not assessed at follow up because they were too unwell, declined to be interviewed, or were not contactable. In these CHR participants, transition or non-transition to psychosis was determined from their clinical records, but it was not possible to assess their level of functioning.

### MRI data acquisition and preprocessing

The present study analysed task-based fMRI data and ^1^H-MRS data acquired from the same participants during the same MRI session. Details of the fMRI novelty salience task used, scan acquisition parameters, preprocessing, modelling of fMRI data, and the acquisition and analysis of hippocampal ^1^H-MRS data have previously been described in detail in publications that report each data modality separately ^13,15^ (Supplementary figure 1). We include the MRS scan quality parameters for the current cohort in Supplementary tables 1, 2 and 3. In the present study, fMRI data analysis focused on the task contrast of novel > neutral oddball trials during the novelty salience task, which was used as a measure of ‘pure stimulus novelty’ ^13 22^.

### Statistical analysis

#### Demographic and behavioural data

Differences between HC and CHR participants for age, gender and handedness were assessed using independent sample *t*-tests (for continuous data) or chi-square (for categorical data) in SPSS 23 (https://www.ibm.com/uk-en/products/spss-statistics). Alongside group comparisons of all CHR vs. HC participants, the CHR group was subdivided according to clinical and functional outcomes at follow-up. Clinical outcome groups (transition to psychosis) comprised CHR participants that had developed psychosis (CHR-T) and those that had not (CHR-NT). Functional outcome was defined as the GAF score at the end of the follow up period, with a score > 65 corresponding to a good level of functioning (CHR-good), and a score of <65 corresponding to a poor level of functioning (CHR-poor) ^15,23^. Group differences during the fMRI novelty salience task, for reaction time, target recognition and error rate were assessed using independent sample *t*-tests in SPSS 23. Significant results are reported at *p* <.05.

### fMRI & ^1^H-MRS data analysis

#### Group x ^1^H-MRS interaction (effects during novel > neutral oddball trial)

To test our a-priori hypothesis, we used multivariate random effects GLM in SPM 12 (https://www.fil.ion.ucl.ac.uk/spm/software/spm12/). Specifically, we tested interaction effects between Group (CHR-NT vs. CHR-T, CHR-good vs. CHR-poor, and HC vs. CHR) and ^1^H-MRS Glx metabolite concentrations on hippocampal functional activity during novel>neutral oddball trials. We chose to use hippocampal Glx metabolite concentrations (combined glutamate and glutamine) as i) the composite Glx peak has been widely used as a marker of glutamatergic function, because it predominantly reflects glutamate levels, which are typically 5 to 6 times higher than those of glutamine ^24^ and ii) a previous meta-analysis reports robust alterations in ^1^H-MRS Glx metabolite concentrations in schizophrenia ^25^. Given our previous findings in this CHR cohort ^15^ additional interaction analysis using ^1^H-MRS glutamate metabolite concentrations are also reported (see Supplementary table 4).

Current tobacco use and age were included as nuisance covariates, as in previous analyses ^13,15^. We conducted ANOVA using hippocampal Glx metabolite concentration as a covariate of interest, restricting the search area to our a-priori region of interest within the bilateral hippocampus (AAL in WFU Pickatlas toolbox; https://www.nitrc.org/projects/wfu_pickatlas).

The initial alpha was set to .*005*, before applying a small volume correction (SVC) for the hippocampal region-of-interest (ROI) analysis, at a voxel-wise threshold of peak level family-wise error (FWE) *p* <0.017^26^ to correct for 3 group tests, i.e i) CHR-T vs. NT ii) CHR-good vs. CHR-poor and iii) CHR vs. HC.

#### Psychophysiological Interaction

To test our a-priori hypothesis regarding the effects of Group and ^1^H-MRS glutamatergic metabolite concentration on hippocampal – striatal functional connectivity, we used a Psychophysiological Interaction (PPI) analysis^49^. Based on novel>neutral oddball trials we included all subjects who showed significant activity within the hippocampal ROI. First, eigenvariates from the hippocampal seed region were extracted from the subject-specific model of the Group x ^1^H-MRS Glx analyses described above. The subject-specific response peak was required to be within a 6mm radius sphere of the right hippocampal region [x, y, z = 36 - 34 −4], i.e. within the group peak to be included in the PPI analysis. This was the case in 39 subjects (14 HC, 25 CHR). Subsequently, for each subject a PPI regressor was created via deconvolution of the eigenvariate time series by weighting the resultant time series with the task contrast time series (novel>neutral oddball trials), adjusted for the effect of interest, and reconvolved with the hemodynamic response function. The resulting contrast was submitted to second level random effects GLM to test the interaction effect between Group and ^1^H-MRS Glx metabolite concentrations on hippocampal functional connectivity. ROIs were created using an atlas composed of functional subdivisions of the striatum (ventral, associative), which is commonly applied in Positron Emission Tomography (PET) research ^9^.

We investigated functional connectivity between the hippocampus and two striatal ROIs (ventral and associative striatum subdivisions) based on a preclinical model highlighting ventral striatal changes in psychosis ^8,10,27,28^, and on reports of dopaminergic dysregulation in the associative striatum in psychosis ^29^. The SVC results were considered significant at alpha = .*005* and peak level FWE *p* (*p*_*peak*FWE_) < .025 to adjust for the two striatal ROI (ventral and associative striatum) comparisons.

## Results

### Demographic, clinical and medication data

All demographic, clinical and medication data categorised by group are summarised in Table 1. At baseline the CHR group were younger and smoked more cigarettes, had higher levels of anxiety (HAM-A scores) and depression (HAM-D scores), and a lower level of functioning (GAF scores) than HC. At clinical follow-up, 12 CHR individuals (16% of the total CHR sample) had transitioned to psychosis (CHR-T) and 63 (84%) had not (CHR-NT). At baseline, the CHR-T group smoked fewer cigarettes than the CHR-NT group but did not differ on any other measures (see Table 1). At follow-up, GAF scores were available in 52 CHR participants. Seventeen CHR participants had a follow-up GAF score > 65 indicating a good functional outcome, and 35 had a GAF score < 65 indicating a poor functional outcome. The functional outcome group did not differ on any demographic measure (see Table 1). CAARMS and GAF score significantly correlated (n=45, *p*<0.001, *r*=-0.49), which was expected.

### Behavioural data

During the novelty salience task, the mean reaction time for responses to target stimuli was 544ms (SD=142ms), and the mean number of errors was 1.69 (SD=3.1). There were no significant group differences in mean reaction time or target recognition time for any comparison (CHR-NT vs. CHR-T, CHR-good vs. CHR-poor, HC vs. CHR).

### MRI: Interactions between Group, hippocampal Glx and functional activity

All Group x fMRI x ^1^H-MRS Glx interaction results are summarised in Table 2.

**Table 2:** results split by modality, comparison and lateralisation with the cluster selected for PPI analysis shaded.

| Modality | Comparison | Significant result |
| --- | --- | --- |
| fMRI x MRS<br>Glx | HC > CHR | **Hippocampus ROI**<br>**Right hippocampus**<br>$p_{peakFWE} = .052$, x y z=34 -34 -6, $t=3.46$, $z=3.35$, cluster extent=146 |
| | HC + CHR-NT > CHR-T | **Hippocampus ROI**<br>**Right hippocampus**<br>$p_{peakFWE} = .002$, x y z=36 -34 -4, $t=4.46$, $z=4.24$, cluster extent=140 |
| | CHR-NT > CHR-T | **Hippocampus ROI**<br>**Right hippocampus** [PPI cluster]<br>$p_{peakFWE} = .003$, x y z=36 -34 -4, $t=4.4$, $z=4.19$, cluster extent=93 |
| | CHR-good > CHR-poor | **Hippocampus ROI**<br>**Right hippocampus**<br>$p_{peakFWE} < .001$, x y z=30 -12 -16, $t=5.52$, $z=4.81$, cluster extent=507<br>**Left hippocampus**<br>$p_{peakFWE} < .001$, x y z=-26 -28 -12, $t=5.25$, $z=4.62$, cluster extent=585 |
| PPI | HC > CHR | **Hippocampus x ventral striatum ROI**<br>**Right caudate**<br>$p_{peakFWE} = .016$, x y z = 6 12 0, $t=3.73$, $z=3.39$, cluster extent = 8<br>**Left caudate**<br>$p_{peakFWE} = .014$, x y z = -10 12 -2, $t=3.78$, $z=3.42$, cluster extent = 50 |
| | | **Hippocampus x associative striatum ROI**<br>**Right caudate**<br>$p_{peakFWE} = .011$, x y z = 18 -4 18, $t=4.45$, $z=3.91$, cluster extent = 570<br>**Left caudate**<br>$p_{peakFWE} = .003$, x y z = -20 12 12, $t=4.92$, $z=4.23$, cluster extent = 352 |

#### Transition to psychosis

There was a significant interaction between group (CHR-T vs. CHR-NT) and ^1^H-MRS Glx metabolite concentrations on right hippocampal activation (*p*_*peak*FWE_ =.003, x y z = 36 −34 −4, t=4.4, z=4.19, k = 93). In the CHR participants who later developed psychosis, there was a negative association between Glx metabolite concentrations and right hippocampus activity that was not evident in the CHR participants who did not transition to psychosis (Figure 2A and 2D).

**Figure 2:**
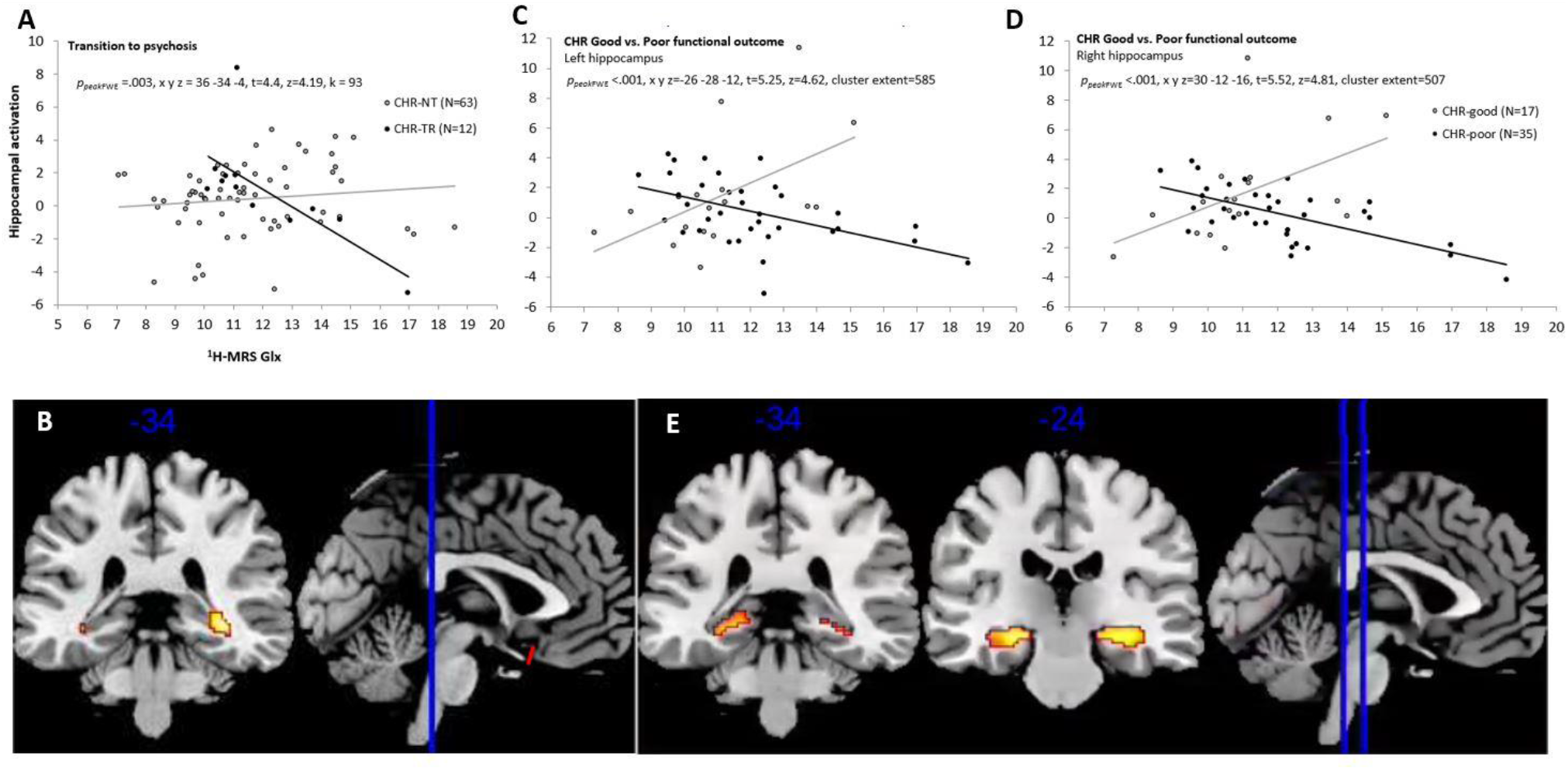
Functional activity. A) Scatterplot showing interaction between Group (CHR-T vs. CHR-NT) x hippocampal Glx on right hippocampal activation during novel > neutral oddball trials. B) SPM brain map (coronal section) showing right hippocampal activation for Group (CHR-T vs. CHR-NT) x hippocampal Glx during novel > neutral oddball trials (P FWE < .017). C & D) Scatterplots showing interaction between Group (CHR-poor vs. CHT-good) x hippocampal Glx on bilateral hippocampal activation during novel > neutral oddball trials. E) SPM brain map (coronal section) showing bilateral hippocampal activation for Group (CHR-good vs. CHR-poor) x hippocampal Glx on novel > neutral oddball trials (pFWE <.017).

#### CHR Good vs. Poor functional outcome

There was a significant interaction between the functional outcome group (CHR-good vs. CHR-poor) and ^1^H-MRS Glx metabolite concentrations on hippocampal activity bilaterally (*p*_*peak*FWE_ *<*.001, x, y, z = 30 −12 −16, t = 5.52, z = 4.81, k =507 & *p*_peakFWE_=.001, x, y, z = −26 −28 −12, t=5.25, z=4.62, k =585). In the CHR participants with a poor functional outcome, there was a negative relationship between hippocampal activity and local Glx concentration. Conversely, in the CHR subjects with a good functional outcome, this relationship was positive (figure 2B, 2C and 2E).

#### CHR vs. HC

The interaction between group (HC vs. CHR) and ^1^H-MRS Glx on hippocampal activation during novel>neutral oddball trials within the bilateral hippocampal ROI was non-significant (*p*_*peak*FWE_ =.052, x y z = 34 −34 −6, t=3.46, z=3.35, k = 146).

### Interactions between Group, hippocampal Glx concentrations and hippocampal-striatal connectivity

PPI analyses focused on a right hippocampal seed region identified by the CHR-NT vs. CHR-T x ^1^H-MRS Glx interaction reported above (extracted eigenvariates x y z = 36 −34 −4). Analysis in relation to transition to psychosis and functional outcome did not reveal any significant PPI effects for hippocampo-striatal connectivity in either ventral or associative striatum ROIs (p > .025 for all analyses). Analysis comparing all CHR vs. HC participants, did reveal a significant interaction effect between group and Glx concentration within the a-priori ventral striatum ROI in the bilateral ventral caudate (*p*_*peak*FWE_ *=*.016, x y z = 6 12 0, t=3.73, z=3.39, cluster extent = 8 and *p*_*peak*FWE_ *=*.014, x y z = −10 12 −2, t = 3.78, z = 3.42, k = 50). An interaction was also observed in the a-priori associative striatum ROI in the dorsal bilateral caudate (*p*_*peak*FWE_ *=*.011, x y z = 18 −4 18, t=4.45, z=3.91, cluster extent = 570 and *p*_*peak*FWE_ *=*.003, x y z = −20 12 12, t = 4.92, z = 4.23, k = 352). In all these regions HC showed a positive association between hippocampal-striatal functional connectivity and hippocampal Glx metabolite concentrations. Conversely, in CHR participants, this relationship was negative (Figure 3).

**Figure 3:**
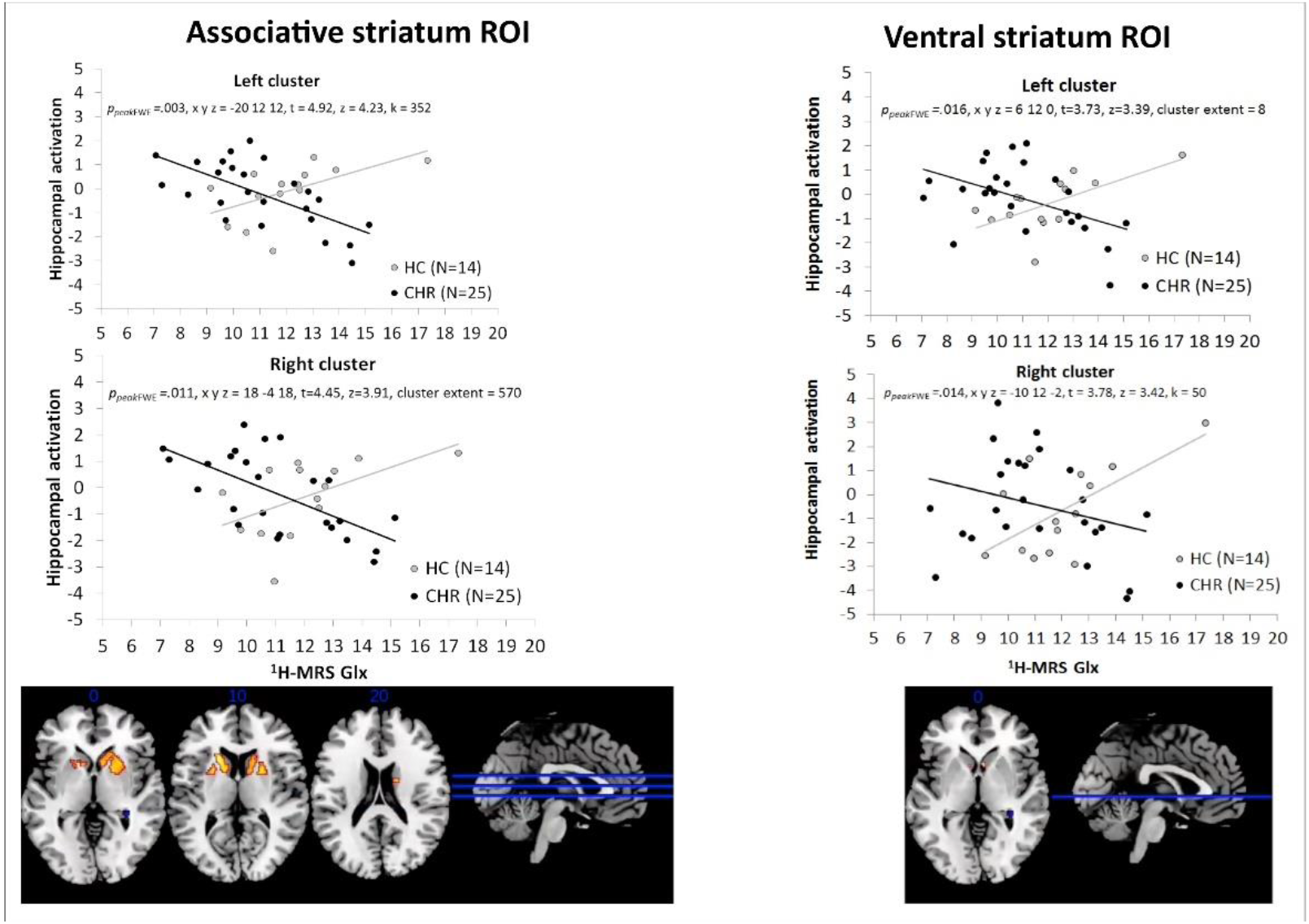
Psychophysiological Interaction. A) Scatterplots showing PPI interactions between Group (CHR vs. HC) in the bilateral associative striatum and B) SPM brain map (axial section) showing bilateral functional connectivity in bilateral associative striatum for the Group (CHR vs HC) x hippocampal Glx during novel > neutral oddball trials (p FEW < .025). C) Scatterplots showing PPI interactions between Group (CHR vs. HC) in the bilateral ventral striatum and B) SPM brain map (axial section) showing bilateral functional connectivity in bilateral ventral striatum for the Group (CHR vs HC) x hippocampal Glx during novel > neutral oddball trails (p FEW < .025).

## Discussion

Preclinical studies indicate that psychosis is associated with increased resting hippocampal activity ^4–6^, and altered hippocampal glutamate activity^5,7^ which is thought to drive an increase in subcortical dopamine activity through glutamatergic projections from hippocampus to striatum ^6^. In previous studies of CHR participants, we reported reduced hippocampal activity during a novelty salience task ^13^, and increased hippocampal glutamatergic metabolite concentrations related to clinical outcomes ^15^. The present study extends these findings and provides support for preclinical models of psychosis by demonstrating that clinical outcomes in CHR participants are also associated with altered *interactions* between hippocampal Glx metabolite concentrations and task-related hippocampal activity. These findings are the first in humans to show that hippocampal glutamatergic concentrations are associated with altered hippocampal activity and function ^7^ in CHR individuals with adverse clinical and functional outcomes. Also in line with preclinical models, our data indicates that the relationship between hippocampal Glx concentrations and hippocampal-striatal connectivity was different in CHR subjects to that of controls, although this association did not influence clinical outcomes.

In the subgroup of CHR subjects that later became psychotic, there was a negative association between hippocampal Glx and hippocampal activation. Similarly, there was a negative association between these measures in CHR subjects who had a low level of functioning at follow-up. A link between this negative association and adverse outcomes is in line with our previous findings from single modality analyses: in separate studies, adverse outcomes were linked to increased hippocampal glutamate metabolite concentrations ^15^ and to reduced hippocampal activation ^13^. To our knowledge, our study presents the first human data to support pre-clinical evidence ^4–7^ that the altered hippocampal activity that predates psychosis reflects dysregulation of local glutamatergic transmission. As discussed by Modinos et al. (2020) the reduced hippocampal activation we observed during novel>neutral oddball stimuli in CHR participants likely reflects a ceiling effect due to increased resting hippocampal activity in CHR subjects ^8,10,27^. Indeed, this was confirmed by our supplementary analysis of Group x ^1^H-MRS interaction effects which showed a *positive* association between hippocampal glutamatergic metabolite concentrations and activity during neutral > standard oddball stimuli in relation to functional outcome (see Supplementary table 5).

In contrast to single modality findings within the CHR group in relation to clinical outcome, we did not find significant differences between the whole CHR group and healthy controls. This is unlikely to reflect a lack of statistical power, as the significant effects relating to outcomes involved CHR subgroups with smaller sample sizes. Rather, it may be related to the heterogeneity of neurobiological findings in the total CHR population: previous neuroimaging studies have reported greater differences within CHR samples in relation to outcomes than between CHR subjects and controls ^15, 30-32^.

Our prediction that the relationship between hippocampal glutamatergic metabolite concentrations and hippocampo-striatal connectivity would be linked to clinical outcomes was not confirmed, although there was an effect in relation to transition that did not survive correction for multiple testing. We examined hippocampal connectivity with the associative (dorsal) striatum due to its role in psychosis and psychosis risk ^31,33–35^, and the ventral striatum as hippocampal glutamatergic outputs project to this sub region ^4,6,29^. There is also evidence for altered salience and reward processing associated with ventral striatal activity in CHR subjects ^11,12,36^. In the present study, we found that the concentrations of Glx in the hippocampus modulated hippocampal functional connectivity with both the ventral and the dorsal caudate, although this effect was distributed more widely in its dorsal portion. For both sub-regions, there was a positive association between hippocampal Glx concentrations and functional connectivity in controls, but the opposite association in CHR subjects. Our data thus provides evidence for an association between both the ventral and the dorsal striatum and increased vulnerability for psychosis.

The study has some limitations. Although the total number of CHR participants that we examined was relatively large for a multi-modal neuroimaging study, because only a minority developed psychosis subsequent to scanning, the size of this subgroup (N=12) was modest. We cannot therefore exclude the possibility that we were unable to detect some differences related to transition to psychosis due to limited statistical power. This issue may be addressed by conducting studies that involve a large number of different centres, permitting the recruitment of larger CHR samples. A more general limitation is that conventional ^1^H-MRS can provide a measure of the mean concentration of glutamatergic metabolites in a given region, but cannot determine which glutamatergic synapses or receptors are involved, or whether the signal reflects the transmitter or metabolic glutamate pools. These issues may be addressed through the development of SPECT or PET ligands that are specific for particular glutamate receptors ^37–39^, and the use of novel MRS techniques ^40^.

Our findings have potential clinical implications. First, they suggest that the ability of tools that are designed to help predict clinical outcomes in CHR subjects may be improved by using multiple, as opposed to single measures ^41,42^. Secondly, they add to existing evidence that hippocampal activity and glutamate function represent promising targets for the development of novel treatments for psychosis^6,43–48^.

In summary, we present the first evidence in humans, in line with rodent models of psychosis, that hippocampal activity and glutamatergic function are associated and that their interactions predict clinical and functional outcomes in CHR subjects. Future research should improve the prediction of outcomes in CHR subjects by incorporating multiple imaging measures in the predictive model, rather than using single risk factors alone.

## Data Availability

Data are available on request.

## Funding

This study was funded by Wellcome Trust grant 091667/Z/10/Z, and supported by the National Institute for Health Research (NIHR) Biomedical Research Centre at South London, Maudsley NHS Foundation Trust and King’s College London. EJH is funded by the Medical Research Council (Grant No. MC- A656-5QD30, Grant No. MC_PC_17214) GM is funded by a Sir Henry Dale Fellowship jointly funded by the Wellcome Trust and the Royal Society (#202397/Z/16/Z). AG is funded by USPHS MH57440. MB was supported by a Veni fellowship from the Netherlands Organization for Scientific Research. AG receives research funding from Alkermes, Lundbeck, and receives consulting fees from Takeda, Roche, Lyra, and Concert. ODH has received investigator-initiated research funding from and/ or participated in advisory/speaker meetings organized by Astra-Zeneca, Autifony, BMS, Eli Lilly, Heptares, Jansenn, Lundbeck, Lyden-Delta, Otsuka, Servier, Sunovion, Rand and Roche. Neither ODH or his family have been employed by or have holdings/a financial stake in any biomedical company.

## Notes

### Competing Interest Statement

The authors have declared no competing interest.

### Clinical Trial

This was not a clinical trial.

### Author Declarations

The study was approved by the National Research Ethics Service Committee of London-Camberwell St Giles, United Kingdom.

